# Assessing the Ability of GPT to Generate Illness Scripts: An Evaluation Study

**DOI:** 10.1101/2023.12.25.23300525

**Authors:** Yasutaka Yanagita, Daiki Yokokawa, Fumitoshi Fukuzawa, Shun Uchida, Takanori Uehara, Masatomi Ikusaka

## Abstract

**Background:** Illness scripts, which are structured summaries of clinical knowledge concerning diseases, are crucial in disease prediction and problem representation during clinical reasoning. Clinicians iteratively enhance their illness scripts through their clinical practice. Because illness scripts are unique to each physician, no systematic summary of specific examples of illness scripts has been reported.

**Objective:** Generative artificial intelligence (AI) stands out as an educational aid in continuing medical education. The effortless creation of a typical illness script by generative AI could enhance the comprehension of disease concepts and increase diagnostic accuracy. This study investigated whether generative AI possesses the capability to generate illness scripts.

**Methods:** We used ChatGPT, a generative AI, to create illness scripts for 184 diseases based on the diseases and conditions integral to the National Model Core Curriculum for undergraduate medical education (2022 revised edition) and primary care specialist training in Japan. Three physicians applied a three-tier grading scale: “A” if the content of each disease’s illness script proves sufficient for training medical students, “B” if it is partially lacking but acceptable, and “C” if it is deficient in multiple respects. Moreover, any identified deficiencies in the illness scripts were discussed during the evaluation process.

**Results:** Leveraging ChatGPT, we successfully generated each component of the illness script for 184 diseases without any omission. The illness scripts received “A,” “B,” and “C” ratings of 56.0% (103/184), 28.3% (52/184), and 15.8% (29/184), respectively.

**Conclusion:** Useful illness scripts were seamlessly and instantaneously created by ChatGPT using prompts appropriate for medical students. The technology-driven illness script is a valuable tool for introducing medical students to disease conceptualization.

## Introduction

The illness script encompasses key elements of diseases such as pathophysiology, epidemiology, time course, symptoms and signs, diagnosis, and treatment.^1^ The scripts are systematic summaries of a clinician’s knowledge about a disease and these are associated with problem representations, serving as memorization aids in clinical reasoning. ^2, 3^ Notably, reports suggest that leveraging illness scripts can improve the instruction of clinical reasoning, and serve as an effective method for refining the learner’s clinical reasoning skills. ^4–6^ Therefore, illness scripts increase diagnostic accuracy and are useful for continuing medical education. ^7^

Conversely, the clinical application of illness scripts is not straightforward. Clinicians iteratively enhance their illness scripts through their clinical practice and by encountering a spectrum of cases, including those considered atypical. Illness scripts are not static; they refine and develop as clinicians enhance their skills. Therefore, no standardized illness scripts exist for any disease, and creating them is time-consuming. Hence, our focus on large language models (LLMs), is occasioned by the notable progress achieved in natural language processing using generative pretrained transformers (GPT).^8^ Generative AI, with its potential to function as a virtual educational assistant, stands out in providing information relevant to medical students.^9, 10^ Although generative AI, as typified by ChatGPT, was not explicitly designed for medical applications, previous research has showcased ChatGPT’s capability to successfully pass medical licensing examinations in the United States and Japan.^11, 12^ It has contributed to generating differential diagnosis lists from patient histories,^13^ diagnosed rare diseases,^14^ and intervened in various aspects of medicine. Furthermore, the potential of AI models in specialized medical education and practice is acknowledged.^15^ Therefore, we considered that using generative AI tools such as ChatGPT to generate basic illness scripts holds the potential for physicians to facilitate an effortless grasp of disease concepts by gaining familiarity with illness scripts.

No research has delved into the automated generation of illness scripts tailored to individual diseases. Furthermore, when integrating such technologies into the medical domain, the output’s accuracy becomes critical due to the implications for disease diagnosis and treatment. Because ChatGPT is known to output incorrect information, in this study, board-certified physicians assessed whether ChatGPT can adeptly generate an illness script containing adequate information for physicians to conceptualize the disease.

## Methods

### Study Design

Focusing on the illnesses and conditions integral to the National Model Core Curriculum for undergraduate medical education (2022 revised edition)^16^ and primary care training program in Japan, illness scripts for 184 diseases were systematically generated using ChatGPT. Subsequently, three board-certified physicians conducted an evaluation to gauge the utility of the generated output reached the level required for graduating medical students. of the generated output for medical students. Finally, each illness script was subjected to a comprehensive grading on a three-point scale: “A”: the content proved sufficient for medical students, “B”: it exhibited partial inadequacy, and “C”: it was deemed inadequate in multiple aspects.

### Large language model environment

The illness scripts were generated on July 25, 2023, using the July 20 version of GPT-4 (OpenAI, San Francisco, California, USA). GPT is a large language model (LLM) developed by OpenAI for natural language processing. Its dynamic response generation is based on probabilities the neural network derives from learned syntactic and semantic relationships in text.^17^

### Selecting diseases for illness scripts

Commonly and frequently encountered diseases were selected owing to their importance for medical students. Considering that the diseases managed in primary care overlap with those that medical students should learn about, the diseases studied in primary care training in Japan^18^ were used as a reference. Of the 205 disease and symptom items representing the 16 areas targeted for appropriate management in primary care^18^, 184 were identified as sufficiently relevant for the creation of the illness script. These diseases are also included in the National Model Core Curriculum in Japan for undergraduate medical education (2022 revised edition)^16^.

The three board-certified physicians established the exclusion criteria through collaborative discussions and excluded 21 items with minimal diagnostic contribution or mere symptomatology. Seventeen items, e.g., those associated with palliative care or non-critical symptoms (such as lower back pain) were omitted because they lacked the specificity for script creation. Furthermore, four items related to community-acquired pneumonia, herpes encephalitis, herpes infections, and adrenal insufficiency were excluded because they were pertinent to the input examples in the prompt. The English names for the 184 selected items were entered into the prompt based on the International Classification of Diseases, 11th Revision (ICD-11)^19^ registered disease names entered into the prompt for reference (Supplementary Material).

### Content to be entered into ChatGPT, program code

The prompts for ChatGPT were meticulously engineered to ensure their interpretability by generative AI while succinctly defining the desired outputs.^20^ The output items referencing the proposed elements of illness scripts, were determined after discussions facilitated by two board-certified physicians (YY and DY).^1^ The input-specified key elements of the illness scripts included pathophysiology, epidemiology, time course, signs and symptoms, diagnosis, and treatment. The character limit per item was set at less than 50 characters, based on findings from prior illness scripts^1^ and the general requirement that an average of 20-30 words per English sentence could be output. Three output examples (Community-acquired pneumonia, Herpes Zoster, Primary adrenal insufficiency) were added after key elements. The structured prompt for ChatGPT was: [Create an illness script for <DISEASE name>. List the following items in less than 50 characters each: [pathophysiology][epidemiology][time course][Symptoms and Signs][Diagnostics][and treatment]. The following is a reference example of an illness script. Example1), Example2), Example3)] (Figure 1).

**Figure 1:**
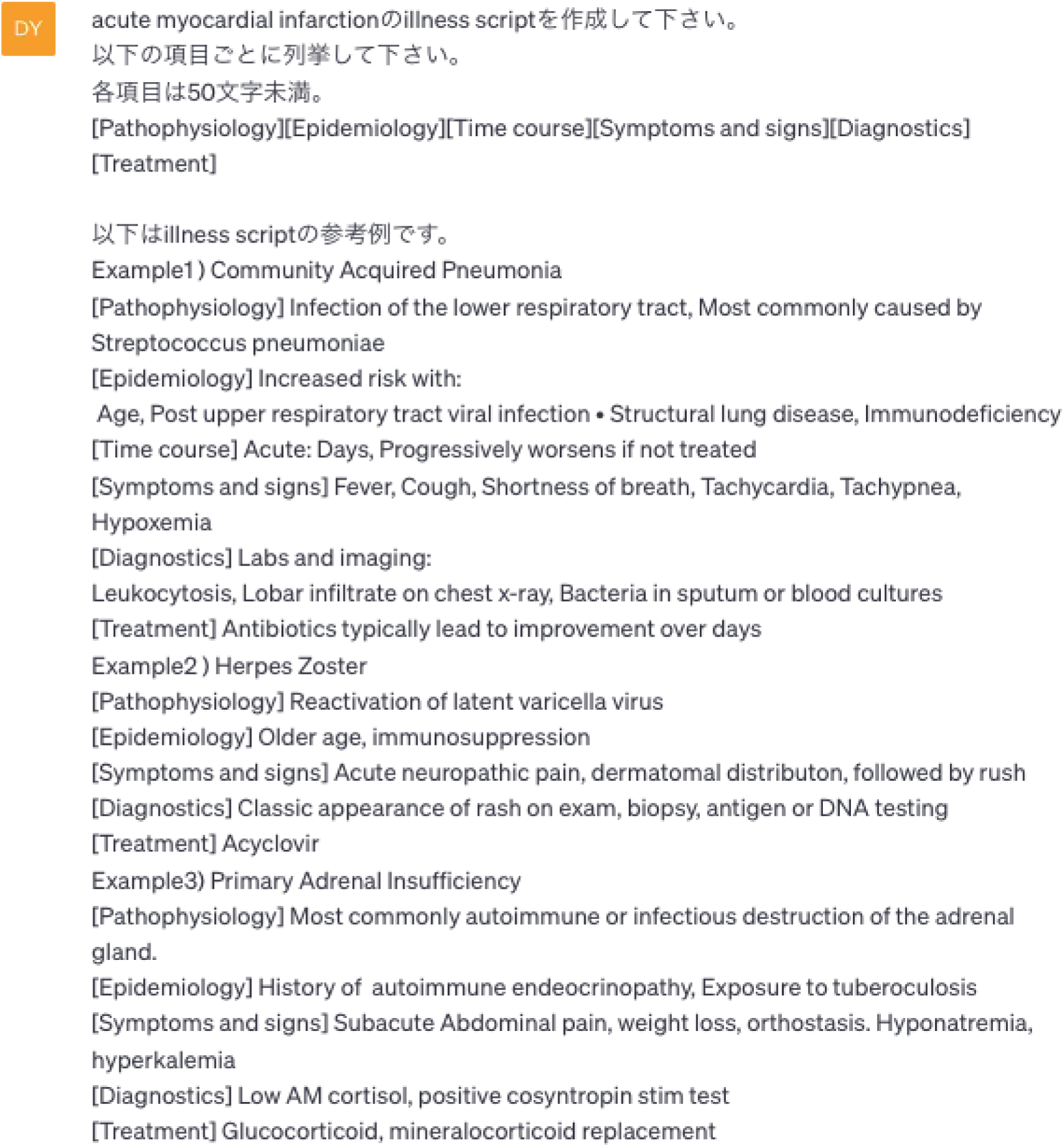
Screenshot of prompt input.

### Evaluation

A broader evaluation was conducted by one board-certified physician and two family physicians (YY, SU, and FF) to assess the generated illness script’s utility for novices, specifically, medical students. Validation involved a comprehensive review by a board-certified physician (YY) to ensure that the output encompassed the essential elements of the illness script: pathophysiology, epidemiology, time course, symptoms and signs, diagnosis, and treatment. The evaluation was structured on a five-point scale, where 1 denoted “not at all useful, needs overall revision,” and 5 represented “very useful, no additional modifications needed.” Composite scores were categorized into three levels: 15, 14, and 13 or less, corresponding to “A,” “B,” and “C,” respectively. Moreover, any identified deficiencies in the illness scripts were discussed during the evaluation. Consequently, an “A” rating signified a script that proved sufficiently informative for medical students and required no further modification, “B” indicated a script that was partially sufficient or required minor revision but was acceptable. “C” represented a script that was inadequate in several respects and necessitated multiple revisions.

### Ethical considerations

This study did not involve human or animal participants, thereby obviating the need for ethical approval.

## Results

Illness scripts for all 184 target diseases were successfully generated using ChatGPT. Each script comprehensively incorporated the specified elements: pathophysiology, epidemiology, time course, signs and symptoms, diagnosis, and treatment, strictly following the prompt instructions. The distribution of the ratings among the evaluated scripts revealed that 56.0% (103/184) were rated “A,” 28.3% (52/184) “B,” and 15.8% (29/184) “C”. Examples of the illness scripts with “A” and “C” ratings are presented in Figures 2 and 3, respectively. The breakdown of evaluations across different categories is provided in Table 1. Cardiovascular and psychiatric systems had the highest number of “C” ratings.

**Figure 2:**
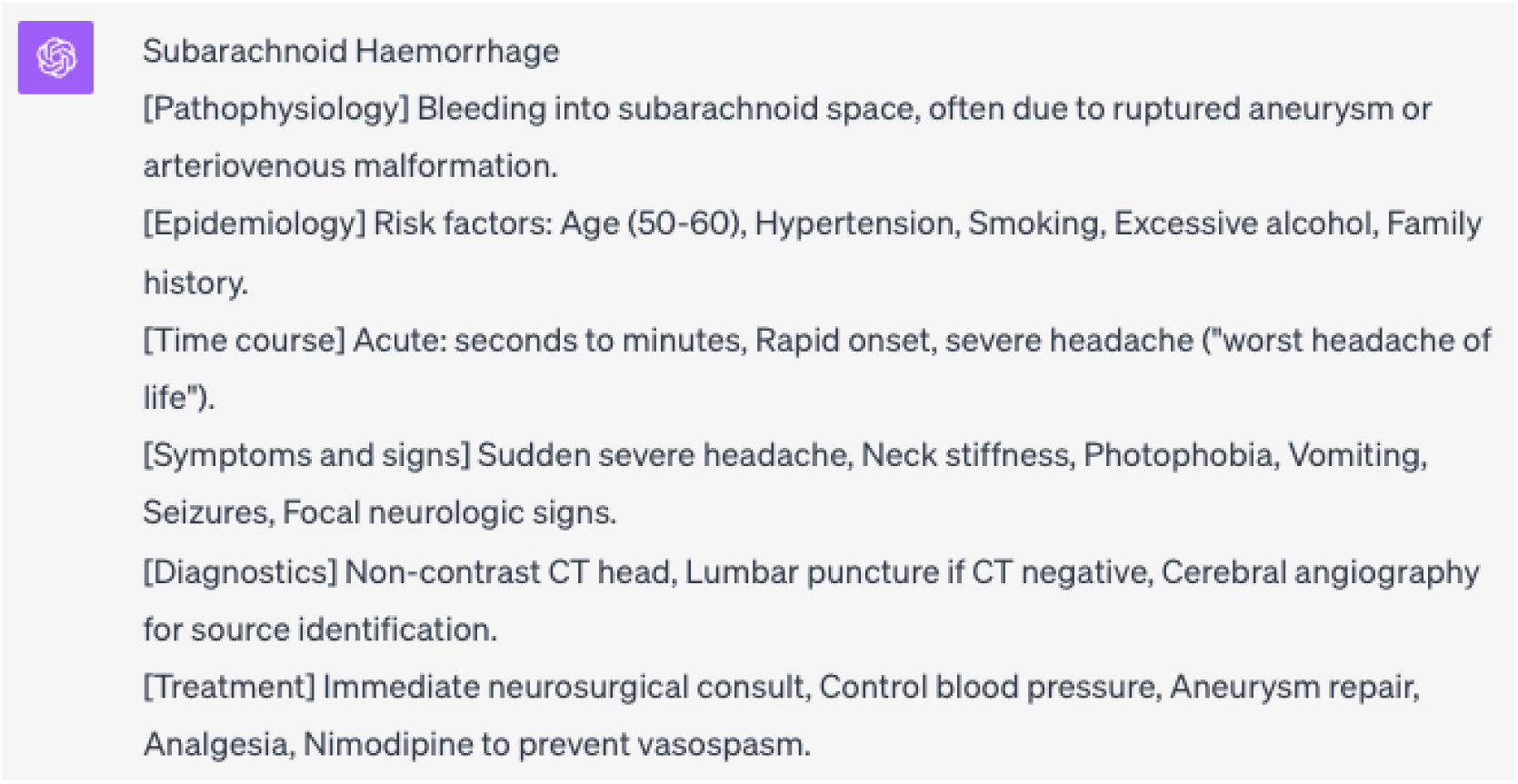
Example of an A-rated illness script.

**Figure 3:**
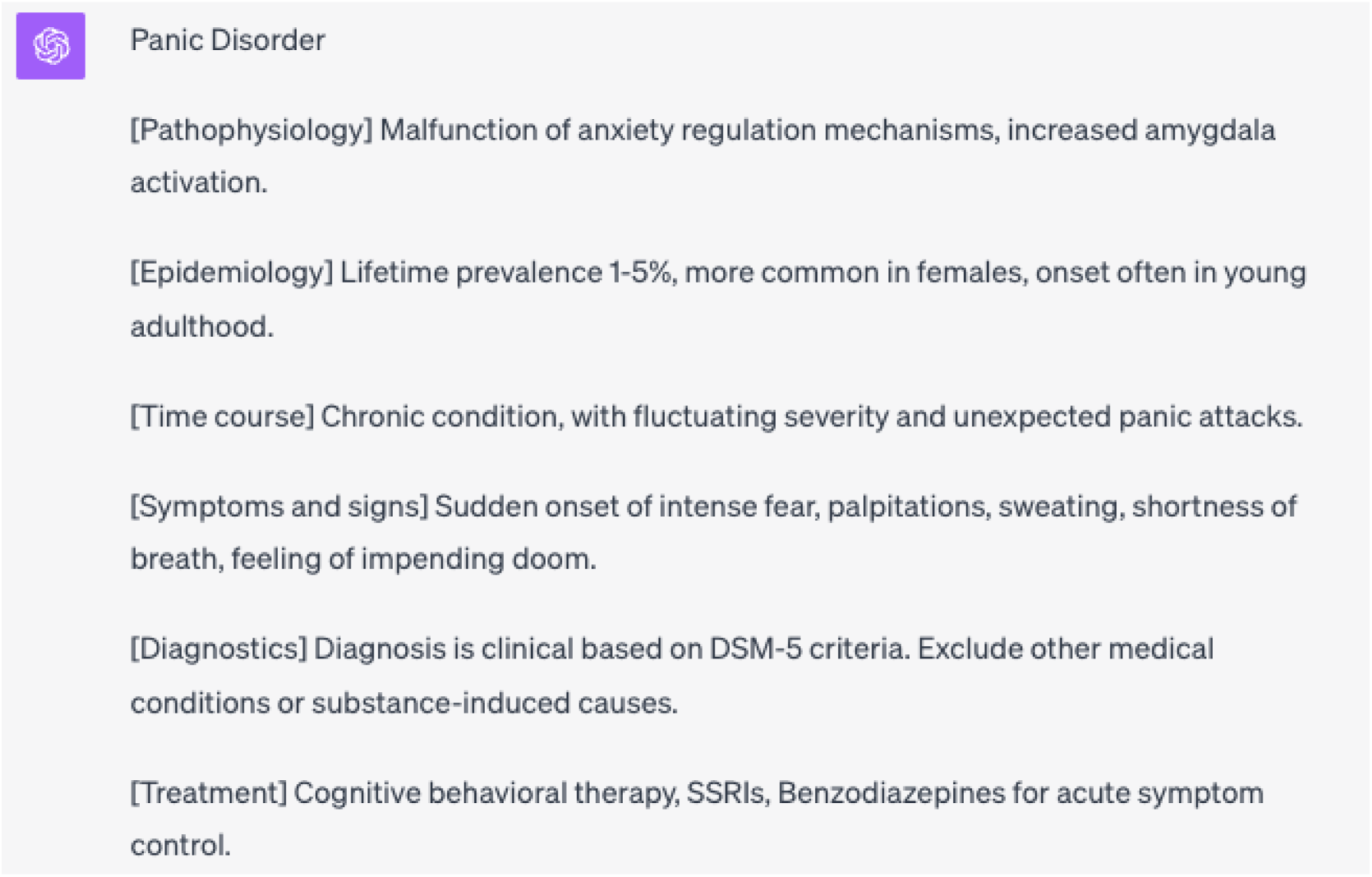
Example of a C-rated illness script.

**Table 1.** Distribution of ratings by 16 areas.

| Classification | Rate A | Rate B | Rate C | Total |
| --- | --- | --- | --- | --- |
|  | (n (%)) | (n (%)) | (n (%)) |  |
| Hematological system | 1 (20.0) | 3 (60.0) | 1 (20.0) | 5 |
| Neurological system | 2 (20.0) | 5 (50.0) | 3 (30.0) | 10 |
| Dermatological system | 8 (61.5) | 5 (38.5) | 0 (0.0) | 13 |
| Musculoskeletal System | 8 (80.0) | 1 (10.0) | 1 (10.0) | 10 |
| Cardiovascular System | 6 (42.9) | 2 (14.3) | 6 (42.9) | 14 |
| Respiratory System | 8 (72.7) | 3 (27.3) | 0 (0.0) | 11 |
| Gastrointestinal System | 17 (68.0) | 7 (28.0) | 1 (4.0) | 25 |
| Renal and Urinary system | 3 (33.3) | 5 (55.6) | 1 (11.1) | 9 |
| Pregnancy and Reproductive System | 10 (66.7) | 3 (20.0) | 2 (13.3) | 15 |
| Endocrine, Nutritional, and<br>Metabolic System | 2 (25.0) | 4 (50.0) | 2 (25.0) | 8 |
| Ophthalmological and Visual System | 6 (75.0) | 1 (12.5) | 1 (12.5) | 8 |
| Otorhinolaryngological and Oral<br>Cavity | 9 (90.0) | 0 (0.0) | 1 (10.0) | 10 |
| Psychiatric System | 4 (36.4) | 2 (18.2) | 5 (45.5) | 11 |
| Infectious | 3 (30.0) | 4 (40.0) | 3 (30.0) | 10 |
| immunologic and Allergic | 1 (50.0) | 1 (50.0) | 0 (0.0) | 2 |
| Physical and Chemical Factors | 3 (50.0) | 3 (50.0) | 0 (0.0) | 6 |
| Pediatric | 10 (66.7) | 4 (26.7) | 1 (6.7) | 15 |
| Geriatrics | 2 (100.0) | 0 (0.0) | 0 (0.0) | 2 |
| Total | 103 (56.0) | 52 (28.3) | 29 (15.8) | 184 |

Deficiencies in the output of ChatGPT’s illness scripts were identified during a comprehensive discussion among one board-certified physician and two family physicians (YY, SU, and FF), specifically focusing on the deduced points. The deficiencies identified within each component of the illness script are outlined below:

Pathophysiology:

1. Droplet transmission for varicella was incorrectly indicated as a route of infection.

Epidemiology:

1. The phrase “Risk: Age” was unclear regarding the specific age group to which it referred.
2. Genetic diseases, such as von Willebrand disease, lacked the associated family history.
3. Phrases like “more common in certain ethnic groups” were deemed too vague.

Time course:

1. The duration of a single attack for cluster headaches was not mentioned besides the symptomatic period.

Diagnostics:

1. Outputs were criticized for being too generic, such as “refer to guidelines” or “exclude similar conditions.”
2. In mitral valve insufficiency and aortic valve stenosis, “Heart murmur on auscultation” is described but the type of murmur is not described.

Treatment:

1. A paper bag (no longer recommended, especially for adults) was suggested for hyperventilation syndrome.
2. Inappropriate antibiotic treatment was output for non-purulent mastitis.
3. The use of the abbreviation VNS was noted for treating epileptic encephalopathy.
4. The treatment for tension headaches is listed as NSAIDs or triptans, but only “use prophylaxis if the frequency is high.” Limited and contradictory data exist concerning the effectiveness of triptans in treating tension-type headaches.
5. In abdominal aortic aneurysms, the description is “Monitor small aneurysms, surgical repair (open or endovascular) for large or rapidly growing aneurysms.” The definitions of small and large are unclear.

## Discussion

This study used ChatGPT to construct typical and easily comprehensible illness scripts for 184 diseases based on the topics covered in the National Model Core Curriculum for undergraduate medical education (2022 revised edition) and primary care residency programs in Japan. The three physicians assigned an “A” rating to 56% of these illness scripts, signifying their adequacy and comprehensiveness. Over half of the generated illness scripts required no changes. Furthermore, 28.3% were rated “B,” indicating partial sufficiency with potential usability after minor revisions and additions. The “A” and “B” ratings, i.e., approximately 84% of the illness scripts, demonstrated relatively high accuracy.

Illness scripts rated as “B” exhibited specific characteristics, such as omitting family history as a crucial risk factor for genetic diseases. In the case of tension headache, the treatment was only indicated as preventive medication if the frequency was high, suggesting the need for more specificity in the output. However, given that one reviewer found the content partially insufficient while the others deemed it sufficient, the overall content was arguably adequate for medical students. Adjusting the character limit restrictions is a potential solution to address this variability.

Illness scripts receiving a “C” rating lacked critical information for diagnosis, such as missing essential symptoms or tests. The evaluator noted that the valvular disease illness script should describe the presence of a heart murmur and the type of murmur. This assessment points to a potential influence from the learning material on which the generative AI was trained. The information on the web on valvular disease is expected to be described only in the presence or absence of a heart murmur, which may have led to inadequate AI output. Such errors can occur in a certain percentage of outputs when generating large volumes of content.

Notably, the prevalence of “A” and “B” ratings was observed throughout the 16 areas regarding the accuracy of illness scripts across different diseases. However, the cardiovascular and psychiatric scripts exhibited a higher proportion of “C” ratings. A more explicit description was required to treat abdominal aortic aneurysms because of variations in the treatment approaches based on the aneurysm size. In the psychiatric system, outputs such as “Diagnosis based primarily on clinical interview and symptom criteria (DSM-5)” were considered too general and lacking specificity. Constraints on the item’s character count may have contributed to the challenge of providing detailed information, particularly given the multifaceted nature of cardiovascular assessments and the wide variety of psychiatric symptoms. The illness scripts compilations of symptoms and tests may not have been output considering the frequency of symptoms or the sensitivity of the tests.

This study employed a straightforward input approach to ChatGPT, specifying three examples of illness scripts in the prompt to control the output standard. Despite setting character limits for each item to minimize redundant information, several illness scripts lacked essential details. Adjusting the character limits for prompts or prioritizing symptoms based on frequency could improve the output for specific diseases or conditions. Furthermore, modifying the character count may allow more accurate illness scripts to be created, especially for complex systems like the cardiovascular system, which had many “C” ratings.

The illness scripts generated in this study underscore the potential of generative AI to produce medical information rapidly and relatively accurately, which confirms its growing applicability in medical education and healthcare. Although concerns persist regarding copyright issues and the medical accuracy of content generated by these generative AI systems,^21^ careful consideration and appropriate use can significantly expand their utility. Medical educators can curate outputs, thus enabling generative AI to be used supportively in delivering educational information to students. Furthermore, these tools hold promise in aiding clinical diagnosis, with illness scripts that assess whether a patient’s symptoms are consistent with known disease presentations, offering immediate practical utility.

It is anticipated that this technology can be adapted to nursing with relative ease. Nursing, which requires a comprehensive understanding of pathophysiology, diagnosis, treatment, and a broader range of information encompassing management and implications, can benefit from tailored prompts fed into generative AI. Extending the illness script concept^22^ to other potential applications across the healthcare field is presently being researched. AI could play a pivotal role in providing valuable insights and information by generating these extended illness scripts.

### Limitation

This study was subject to three limitations: First, the evaluation was conducted based on the GPT-4 version available from July 25, 2023. Given that further updates are anticipated, continuous evaluation is essential

Second, the absence of clear standards for evaluating illness scripts is noteworthy. This study relied on the subjective assessments of three physicians, and results might vary if evaluated by physicians from other specialties.

Third, the utility was evaluated based on whether fundamental aspects of diseases were output for medical students. The study did not verify the usefulness for educators or specialists in various fields, which represents an avenue for future research.

## Conclusion

Generative AI enables the swift and seamless production of illness scripts that can benefit medical students. While the results must be carefully reviewed, the potential applications of this technology in medical education are evident. AI-generated illness scripts may serve as a foundational resource for medical students and provide the basis for developing and refining their own scripts.

## Authors’ Contributions

YY, DY, and MI designed and coordinated the study. YY, DY, FF, and SU conducted the data analysis and interpretation. YY, DY, and MI drafted the manuscript. SU, FF, and UT revised it for important intellectual content. All authors read and approved the final manuscript and agree to be accountable for all aspects of the work in ensuring that questions related to the accuracy or integrity of any part of the work are appropriately investigated and resolved.

## Acknowledgment

We are deeply grateful to the Department of General Medicine members in the Chiba University Hospital for their support. Neither ChatGPT nor other large language models were used in the preparation of this manuscript.

## Data Availability

Data on the results of this study are available from the corresponding author (YY) upon reasonable request.

## Glossary

AI: Artificial Intelligence
LLM: Large Language Models
GPT: Generative Pretrained Transformer
ICD-11: International Classification of Diseases 11th Revision
DSM-5: Diagnostic and Statistical Manual of Mental Disorders-5

