## Supplementary Material for "Assessing the Ability of GPT to Generate Illness Scripts: An Evaluation Study"

Target diseases

| No | Classification | Disease |
| --- | --- | --- |
| 1 | Hematological system | Iron deficiency anaemia |
| 2 | Hematological system | Acute myeloid leukaemia |
| 3 | Hematological system | Malignant lymphoma |
| 4 | Hematological system | Von Willebrand disease |
| 5 | Hematological system | Immune thrombocytopenic purpura |
| 6 | Neurological system | Cerebral ischaemic stroke |
| 7 | Neurological system | Intracerebral haemorrhage |
| 8 | Neurological system | Subarachnoid haemorrhage |
| 9 | Neurological system | Traumatic epidural haemorrhage |
| 10 | Neurological system | Traumatic subdural haemorrhage |
| 11 | Neurological system | Parkinson disease |
| 12 | Neurological system | Bacterial meningitis |
| 13 | Neurological system | Migraine |
| 14 | Neurological system | Tension-type headache |
| 15 | Neurological system | Cluster headache |
| 16 | Dermatological system | Iirritant contact dermatitis |
| 17 | Dermatological system | Atopic eczema |
| 18 | Dermatological system | Asteatotic eczema |
| 19 | Dermatological system | Acute urticaria |
| 20 | Dermatological system | Exanthematic drug eruption |
| 21 | Dermatological system | Impetigo |
| 22 | Dermatological system | Bacterial cellulitis |
| 23 | Dermatological system | Dermatophytosis of foot |
| 24 | Dermatological system | Candidosis of skin |
| 25 | Dermatological system | Acne |
| 26 | Dermatological system | Infected epidermoid cyst |
| 27 | Dermatological system | Molluscum contagiosum |
| 28 | Dermatological system | Scabies |
| 29 | Musculoskeletal System | Fracture of lumbar spine |
| 30 | Musculoskeletal System | Fracture of neck of femur |
| 31 | Musculoskeletal System | Fracture of lower end of radius |
| 32 | Musculoskeletal System | Osteoarthritis of hip |
| 33 | Musculoskeletal System | Strain or sprain of ankle |
| 34 | Musculoskeletal System | Nursemaid's elbow |
| 35 | Musculoskeletal System | Calcific tendinitis |
| 36 | Musculoskeletal System | Osteoporosis |
| 37 | Musculoskeletal System | Intervertebral disc degeneration of lumbar spine with prolapsed disc |
| 38 | Musculoskeletal System | Spinal stenosis |
| 39 | Cardiovascular System | Congestive heart failure |
| 40 | Cardiovascular System | Angina pectoris |
| 41 | Cardiovascular System | Acute myocardial infarction |
| 42 | Cardiovascular System | Atrial fibrillation |
| 43 | Cardiovascular System | Complete atrioventricular block |
| 44 | Cardiovascular System | Mitral valve insufficiency |
| 45 | Cardiovascular System | Aortic valve stenosis |
| 46 | Cardiovascular System | Lower limb atherosclerosis |
| 47 | Cardiovascular System | Abdominal aortic aneurysm |
| 48 | Cardiovascular System | Deep vein thrombosis |
| 49 | Cardiovascular System | Lower limb varicose veins |
| 50 | Cardiovascular System | Lymphoedema |
| 51 | Cardiovascular System | Essential hypertension |
| 52 | Cardiovascular System | Secondary hypertension |
| 53 | Respiratory System | Acute upper respiratory infections |
| 54 | Respiratory System | Acute bronchitis |
| 55 | Respiratory System | Asthma |
| 56 | Respiratory System | Bronchiectasis |
| 57 | Respiratory System | Chronic obstructive pulmonary disease |
| 58 | Respiratory System | Pneumoconiosis |
| 59 | Respiratory System | Hyperventilation |
| 60 | Respiratory System | Obstructive sleep apnoea |
| 61 | Respiratory System | Pneumothorax |
| 62 | Respiratory System | Pleurisy |
| 63 | Respiratory System | Lung cancer |
| 64 | Gastrointestinal System | Esophageal varices |
| 65 | Gastrointestinal System | Stomach cancer |
| 66 | Gastrointestinal System | Gastric ulcer |
| 67 | Gastrointestinal System | Gastritis |
| 68 | Gastrointestinal System | Gastro-oesophageal reflux disease |
| 69 | Gastrointestinal System | Obstruction of small intestine |
| 70 | Gastrointestinal System | Acute appendicitis |
| 71 | Gastrointestinal System | Haemorrhoids |
| 72 | Gastrointestinal System | Anal fistula |
| 73 | Gastrointestinal System | Irritable bowel syndrome |
| 74 | Gastrointestinal System | Diverticulitis of large intestine |
| 75 | Gastrointestinal System | Cholelithiasis |
| 76 | Gastrointestinal System | Acute cholecystitis |
| 77 | Gastrointestinal System | Cholangitis |
| 78 | Gastrointestinal System | Acute viral hepatitis |
| 79 | Gastrointestinal System | Chronic viral hepatitis |
| 80 | Gastrointestinal System | Hepatic cirrhosis |
| 81 | Gastrointestinal System | Liver cancer |
| 82 | Gastrointestinal System | Alcoholic hepatitis |
| 83 | Gastrointestinal System | Drug-induced liver disease |
| 84 | Gastrointestinal System | Acute pancreatitis |
| 85 | Gastrointestinal System | Chronic pancreatitis |
| 86 | Gastrointestinal System | Acute abdomen |
| 87 | Gastrointestinal System | Diaphragmatic hernia |
| 88 | Gastrointestinal System | Incisional hernia |
| 89 | Renal and Urinary system | Acute kidney failure |
| 90 | Renal and Urinary system | Chronic kidney disease |
| 91 | Renal and Urinary system | Acute nephritic syndrome |
| 92 | Renal and Urinary system | Chronic nephritic syndrome |
| 93 | Renal and Urinary system | Nephrotic syndrome |
| 94 | Renal and Urinary system | Diabetic kidney disease |
| 95 | Renal and Urinary system | Calculus of ureter |
| 96 | Renal and Urinary system | Acute pyelonephritis |
| 97 | Renal and Urinary system | Overactive bladder |
| 98 | Pregnancy and Reproductive System | Normal pregnancy |
| 99 | Pregnancy and Reproductive System | Spontaneous abortion |
| 100 | Pregnancy and Reproductive System | Preterm delivery |
| 101 | Pregnancy and Reproductive System | Spontaneous delivery |
| 102 | Pregnancy and Reproductive System | Obstetric haemorrhage |
| 103 | Pregnancy and Reproductive System | Nonpurulent mastitis |
| 104 | Pregnancy and Reproductive System | Amenorrhoea |
| 105 | Pregnancy and Reproductive System | Abnormal uterine bleeding |
| 106 | Pregnancy and Reproductive System | Menopausal symptom |
| 107 | Pregnancy and Reproductive System | Pelvic inflammatory diseases |
| 108 | Pregnancy and Reproductive System | Ovary tumors |
| 109 | Pregnancy and Reproductive System | Breast tumors |
| 110 | Pregnancy and Reproductive System | Hyperplasia of prostate |
| 111 | Pregnancy and Reproductive System | Erectile dysfunction |
| 112 | Pregnancy and Reproductive System | Testicular tumors |
| 113 | Endocrine and Metabolic System | Acromegaly |
| 114 | Endocrine and Metabolic System | Hyperthyroidism |
| 115 | Endocrine and Metabolic System | Hypothyroidism |
| 116 | Endocrine and Metabolic System | Diabetes mellitus |
| 117 | Endocrine and Metabolic System | Diabetic polyneuropathy |
| 118 | Endocrine and Metabolic System | Diabetic acidosis |
| 119 | Endocrine and Metabolic System | Hypercholesterolaemia |
| 120 | Endocrine and Metabolic System | Gout |
| 121 | Ophthalmological and Visual System | Myopia |
| 122 | Ophthalmological and Visual System | Hyperopia |
| 123 | Ophthalmological and Visual System | Astigmatism |
| 124 | Ophthalmological and Visual System | Allergic conjunctivitis |
| 125 | Ophthalmological and Visual System | Cataracts |
| 126 | Ophthalmological and Visual System | Glaucoma |
| 127 | Ophthalmological and Visual System | Diabetic retinopathy |
| 128 | Ophthalmological and Visual System | Hypertensive retinopathy |
| 129 | Otorhinolaryngological and Oral Cavity | Acute otitis media |
| 130 | Otorhinolaryngological and Oral Cavity | Acute sinusitis |
| 131 | Otorhinolaryngological and Oral Cavity | Chronic rhinosinusitis |
| 132 | Otorhinolaryngological and Oral Cavity | Allergic rhinitis |
| 133 | Otorhinolaryngological and Oral Cavity | Acute tonsilitis |
| 134 | Otorhinolaryngological and Oral Cavity | Foreign body in ear |
| 135 | Otorhinolaryngological and Oral Cavity | Foreign body in nasal sinus |
| 136 | Otorhinolaryngological and Oral Cavity | Foreign body in pharynx |
| 137 | Otorhinolaryngological and Oral Cavity | Foreign body in larynx |
| 138 | Otorhinolaryngological and Oral Cavity | Foreign body in oesophagus |
| 139 | Psychiatric System | Alzheimer disease |
| 140 | Psychiatric System | Dementia due to cerebrovascular disease |
| 141 | Psychiatric System | Alcohol dependence |
| 142 | Psychiatric System | Nicotine dependence |
| 143 | Psychiatric System | Depressive disorder |
| 144 | Psychiatric System | Bipolar type I disorder |
| 145 | Psychiatric System | Schizophrenia |
| 146 | Psychiatric System | Panic disorder |
| 147 | Psychiatric System | Bodily distress disorder |
| 148 | Psychiatric System | Adjustment disorder |
| 149 | Psychiatric System | Insomnia |
| 150 | Infectious | Influenza |
| 151 | Infectious | Measles |
| 152 | Infectious | Rubella |
| 153 | Infectious | Varicella |
| 154 | Infectious | Mumps |
| 155 | Infectious | Human immunodeficiency virus disease |
| 156 | Infectious | Group A streptococcus pyogenes infection |
| 157 | Infectious | Chlamydia infection |
| 158 | Infectious | Tuberculosis |
| 159 | Infectious | Mycetoma |
| 160 | Immunologic and Allergic | Polymyalgia rheumatica |
| 161 | Immunologic and Allergic | Sjögren's syndrome |
| 162 | Physical and Chemical Factors | Alcoholic intoxication |
| 163 | Physical and Chemical Factors | Cocaine intoxication |
| 164 | Physical and Chemical Factors | Anaphylaxis |
| 165 | Physical and Chemical Factors | Heat stroke |
| 166 | Physical and Chemical Factors | Hypothermia |
| 167 | Physical and Chemical Factors | Burns |
| 168 | Pediatric | Epileptic encephalopathies |
| 169 | Pediatric | Pediatric viral infections (measles) |
| 170 | Pediatric | Pediatric viral infections (mumps) |
| 171 | Pediatric | Pediatric viral infections (varicella) |
| 172 | Pediatric | Roseola infantum |
| 173 | Pediatric | Pediatric viral infections (influenza) |
| 174 | Pediatric | Acute bronchiolitis due to respiratory syncytial virus |
| 175 | Pediatric | Gastroenteritis due to Rotavirus |
| 176 | Pediatric | Meningitis due to Staphylococcus |
| 177 | Pediatric | Childhood asthma |
| 178 | Pediatric | Tetralogy of Fallot |
| 179 | Pediatric | Autism spectrum disorder |
| 180 | Pediatric | Learning disorder |
| 181 | Pediatric | Down syndrome |
| 182 | Pediatric | Disorders of intellectual development |
| 183 | Geriatrics | Aspiration in the elderly |
| 184 | Geriatrics | Pressure ulceration in the elderly |

Excluded diseases

| 1 | Musculoskeletal System | Low back pain |
| --- | --- | --- |
| 2 | Cardiovascular System | Cardiomyopathy |
| 3 | Respiratory System | Chronic respiratory failure |
| 4 | Gastrointestinal System | Peritonitis |
| 5 | Renal and Urinary system | Chronic kidney disease, stage 5 |
| 6 | Pregnancy and Reproductive System | Puerperium |
| 7 | Psychiatric System | Psychotic disorders |
| 8 | Infectious | Staphylococcal infection |
| 9 | Infectious | Methicillin resistant staphylococcus aureus |
| 10 | Infectious | Fungal infections |
| 11 | Infectious | Sexually transmitted infections |
| 12 | immunologic and Allergic | Allergic diseases |
| 13 | Geriatrics | Undernutrition |
| 14 | Geriatrics | Falls in the elderly |
| 15 | Geriatrics | Incontinence in the elderly |
| 16 | Malignant tumor | Remission maintenance |
| 17 | Malignant tumor | Palliative care |
| 18 | Neurological system | Encephalitis due to herpes simplex virus |
| 19 | Respiratory System | Bacterial pneumonia |
| 20 | Endocrine and Metabolic System | Adrenocortical insufficiency |
| 21 | Infectious | Herpes |
